# A Deadly Equation: The Global Toll of US TB Funding Cuts

**DOI:** 10.1101/2025.03.04.25323340

**Authors:** Sandip Mandal, Sreenivas Nair, Suvanand Sahu, Lucica Ditiu, Carel Pretorius

## Abstract

**Background:** The abrupt discontinuation of U.S. government funding for global tuberculosis (TB) programs has created significant challenges in TB care and prevention. In 2023, external funding accounted for 21% of total TB expenditures, with the U.S. contributing over 55% of these funds. The withdrawal of U.S. support threatens essential TB services, including diagnostics, treatment, TB-HIV co-infection interventions, and research initiatives critical to achieving the 2030 Sustainable Development Goals (SDGs) for TB eradication.

**Methods:** This study analyses the dependency of 26 high-burden TB countries (HBCs) on U.S. funding and models the potential impact of funding cuts on TB incidence and mortality. Countries were classified into low (0–22%), moderate (23–37%), and high (>37%) dependency groups based on three-year expenditure data. A deterministic compartmental model simulated service disruptions under three recovery scenarios: (1) minimal impact (services recover within three months), (2) moderate impact (recovery within one year), and (3) worst-case scenario (permanent service reduction). Extrapolations were made for all 26 HBCs based on representative countries from each dependency category.

**Findings:** Funding cuts are projected to result in substantial increases in TB cases and deaths across the three scenarios. In a high-dependency country, cumulative TB cases could rise by 2.1% (CI 1.5 – 2.5), 5.4% (CI4 – 6.5), or 36% (CI 25 – 47), and TB deaths by 2.9%(CI 2.2 – 3.5), 7.8%(CI 5.9 – 9.2), 68%(CI 45 - 86), depending on the severity of service disruptions. Across all 26 HBCs, additional TB cases are estimated at 0.63 million (CI 0.45 – 0.81) (minimal impact), 1.66 million (CI 1.2 – 2.1) (moderate impact), and 10.67 million (CI 7.85 – 13.19) (worst-case). Corresponding TB deaths are projected to increase by 99,800 (CI 65,200–130,000), 268,600 (CI 185,800–337,900), and 2,243,700 (CI 1,570,800–2,807,300), respectively. The findings underscore the catastrophic consequences of the funding cut, particularly in countries highly reliant on U.S. support.

**Interpretation:** The loss of U.S. funding endangers global TB control, jeopardizing End TB and SDG targets. While some nations may adapt, short-term disruptions will severely impact vulnerable populations. Urgent alternative funding is needed to sustain TB prevention and treatment efforts and mitigate the effects of aid withdrawal.

**Funding:** None for this specific work

## Introduction

The recent abrupt funding discontinuation by the US Government has impacted TB care and prevention across high TB burden countries^1^. In 2023, total funding for tuberculosis (TB) care and prevention reached US$ 5.7 billion, with 21% (US$ 1.2 billion) coming from external sources. This external funding played a crucial role in sustaining TB programs worldwide. In 2024, USAID committed US$ 406 million (figure S1; appendix p 1)^2^, while the Global Fund (GF) allocated US$ 800 million, of which approximately one-third (∼US$ 267million) originates from the U.S. Government. Consequently, U.S. contributions account for over 55% of the total external funding, highlighting their critical role in the global TB response.^3^

USAID Programs funded by USAID’s TB initiative face disruptions in key areas, including accelerated TB and drug-resistant TB detection and treatment, expanded coverage of interventions for TB-HIV co-infection, prevention and treatment of TB infection, improvements in the TB service delivery platforms, research and innovation—essential to the Global Plan to End TB (2023–2030).^4^ Additionally, critical TB research, essential for developing new tools and achieving the 2030 SDG targets, is under threat. USAID has been a key funder for TB drug and diagnostics development, operational research, and vaccine innovation.^3^ The aid discontinuation jeopardizes these advancements, potentially stalling progress in the fight against TB.

In the face of these challenges, using mathematical modelling, we estimated the potential impact of U.S. TB funding cuts on 26 high-burden countries (HBC). In the following section we describe the basic model framework, the different data sources involved and model scenarios. We present results for projected epidemics on service disruptions under three recovery scenarios: (1) minimal impact, (2) moderate impact, and (3) worst-case scenario. Finally, we discuss implications of this work, the limitations of the model and global relevance.

## Methods

The degree of dependency on US funding for tuberculosis (TB) programs varies widely across countries. Our analysis focusses on 26 high-burden countries (HBC), major recipients of the US international aid, using publicly available expenditure data reported annually by countries to WHO, to estimate an approximate distribution for the proportion of total expenditure that derives directly from US funding.^5^ We used three-year averages expenditure data from 2021-2023 when available and relied on 2023 data if data from 2021 or 2022 were missing.

We categorized these countries into three levels of dependency. Low Dependency (0%–22%): Eight out of 26 HBCs have diversified funding sources with minimal reliance on US aid, making them less susceptible to disruptions in TB services.

Moderate Dependency (23%–37%): Ten countries out of 26 HBCs face moderate risk, where reductions in US funding could strain budgets, affecting service delivery and supply chains. High Dependency (>37%): Eight countries out of 26 HBCs are highly reliant on US funding and are extremely vulnerable to program disruptions, including shortages of diagnostic tools, medications, and healthcare personnel (table S1, appendix pp 1-2). A freeze and discontinuation in aid poses a severe threat to the continuity of TB services in these nations.

## Model

We investigated the potential impact of funding cuts on TB incidence and mortality by modelling service disruptions using, a deterministic, compartmental model of TB transmission under Bayesian framework (figure S2; appendix pp 2-6).^6^ We assumed that the proportion of NTP expenditure funded by USAID will translate proportionately to impact on parameters that influence detection and treatment outcomes. A 10% funding reduction, for instance, would proportionally decrease key probabilities in the TB detection cascade. If the funding gap remains unmet, TB services decline accordingly.

In our analysis, and likely in practice, there is no immediate way to overcome the funding gap and service delivery disruption, sooner than three months after the 90 days evaluation period of the USAID funding freeze. Even in situations where countries can fund the gap themselves, it would take time to restructure procurement and logistics systems. Therefore, we explored three recovery scenarios to estimate the potential impact of a funding freeze and discontinuation.

a. Minimum Impact Scenario (S1): The USAID freeze and funding cut disrupts TB services for 90 days. The funding gap is subsequently met, and the country recovers service coverage to baseline levels within next three months.
b. Moderate Impact Scenario (S2): The USAID freeze and funding cut disrupts TB services for 90 days. The funding gap is subsequently met, and it takes one year for the country to recover service coverage to baseline levels.
c. Worst Possible Scenario (S3): The USAID freeze and funding cut disrupts TB services for 90 days, after which funding gap is not met, and service coverage remains at a lower level permanently.

To better illustrate the potential epidemiological effects of a funding freeze and discontinuation, we choose one country from each dependency category - Country 1: South East Asian country with large TB burden and a high proportion of domestic resources for TB (low dependency); Country 2: South East Asian country with high TB burden with medium domestic resources for TB (moderate dependency); and Country 3: An African country with large TB burden and low domestic resources for TB (high dependency) and calibrated the respective country models with epidemiological data for these countries and produced the three disruption scenarios (table S2, figure S3; appendix pp 5-9).

To estimate the impact for the remaining 23 high-burden countries, we use a simple extrapolation method, avoiding the extensive time required to generate scenarios for each country using calibrated models. Baseline trends for new TB cases and TB deaths were projected using a cubic-spline method, and each has been assigned to the aforementioned-dependency category (low, medium and high).^6^ We then apply the relative impact estimates from the representative country of each category to the baseline trend of the country being extrapolated. Across all scenarios, service disruptions lead to delays in diagnosis and treatment initiation, resulting in substantial increases in TB incidence and mortality (figure S4, appendix p 10).

## Results

In Country 1, the number of additional TB cases from 2025 - 2030 could be increased by 0.5% (CI 0.3% – 0.7%), 1.2%(CI 0.9% – 1.6%) or 6.8%(CI 4.6% – 9.1%) in minimal, moderate or worst possible scenarios respectively. In Country 2 cumulative cases could increase by 1.2%(CI 0.8% – 1.6%), 3.1%(CI 2.2% – 4.3%) or 19%(CI 12.7% – 27.4%) respectively, whereas in Country 3, additional cases could increase by 2.1%(CI 1.5% – 2.5%), 5.4%(CI 4.0% – 6.5%) or 36%(CI 25% - 47%) in the aforementioned scenarios. Simultaneously, the cumulative deaths during this period could increase by 0.8%(CI 0.6% – 1.0%), 2.0%(CI 1.5% – 2.5%) or 13.2%(CI 9.6% – 17.0%) respectively for three scenarios in Country 1. In Country 2, these scenarios project additional TB deaths by 1.9%(CI 1.4% – 2.4%), 4.9%(CI 3.5% – 6.4%) and 37%(CI 25% - 52%) and in Country 3, 2.9%(CI 2.2% – 3.5%), 7.8%(CI 5.9% – 9.2%), 68%(CI 45% - 86%) respectively.

Across the 26 high-burden countries, Scenarios S1-S3 are estimated to result in 634,700 [CI 447,700–806,600], 1,660,000 [CI 1,210,400–2,060,000], and 10,676,400 [CI 7,848,800–13,190,000] additional TB cases, respectively, from 2025 to 2030 (Figure, A and C, also see table S4, appendix p 11). Correspondingly, TB deaths are projected to increase by 99,800 [CI 65,200–130,000], 268,600 [CI 185,800–337,900], and 2,243,700 [CI 1,570,800–2,807,300] across these scenarios during the same period (Figure, B and D, table S4, appendix p 11).

**Figure 1:**
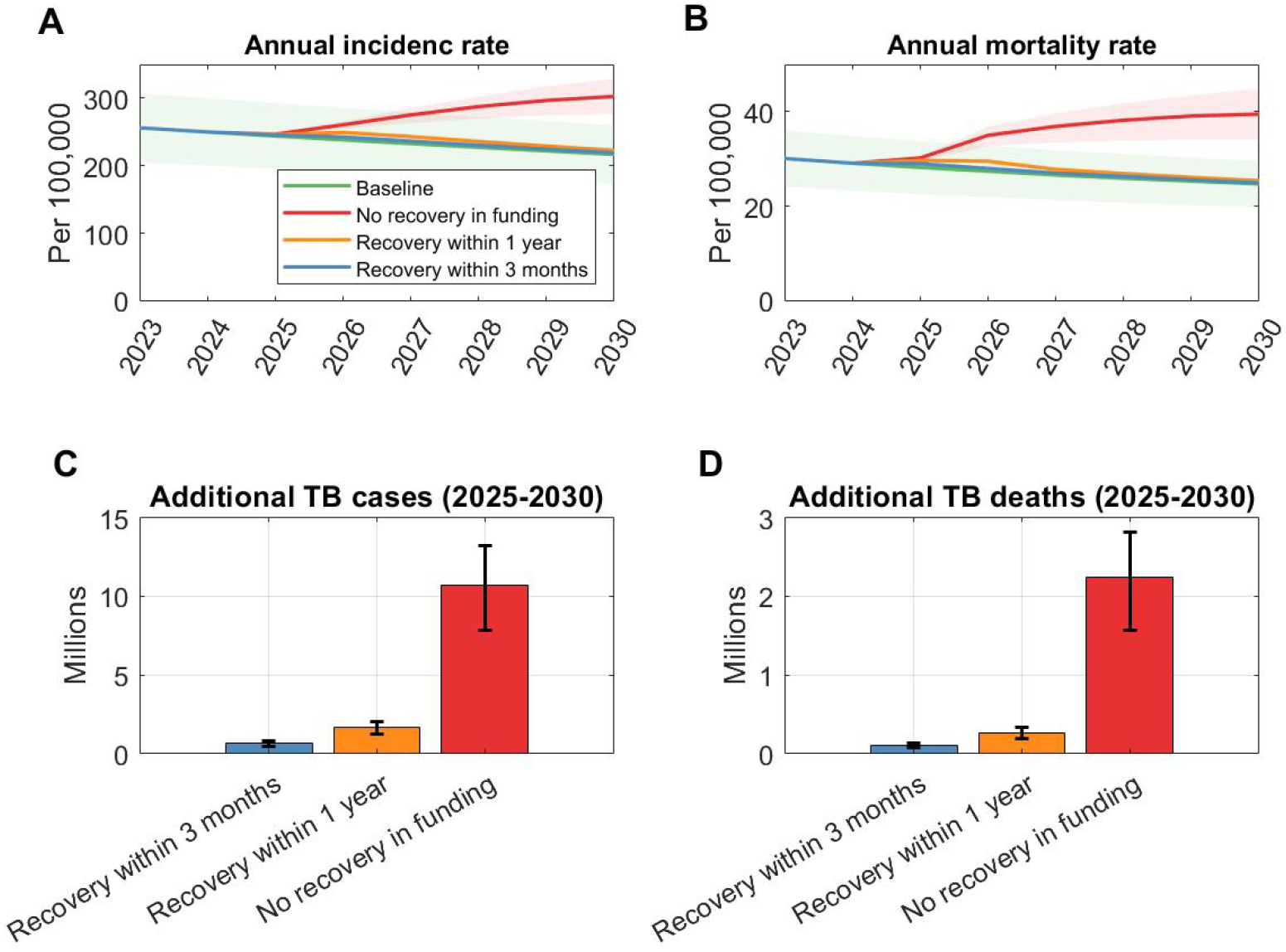
Impact of a US Aid freeze and cut on tuberculosis outcomes across 26 HBCs from 2025 - 2030. (A) Annual TB incidence per 100,000 population under three funding scenarios: a 90-day freeze with a subsequent 3-month recovery, a one-year recovery, and permanent disruption (no recovery). (B) Annual TB mortality rate corresponding to the same three recovery scenarios. (C) and (D) shows additional cases and deaths during 2025 to 2030 under the three recovery scenarios.

Our best estimate is derived from the impact mapping of each country, based on funding contributions reported to WHO. The range of estimates is generated by randomly varying the USAID dependency level assigned to each country – classified as low, medium, or high USAID dependency. This interval is then rescaled so that the median impact level from these variations aligns with the best estimate. This approach serves as a practical method to account for uncertainty in our assumption that a proportional reduction in funding contributions directly corresponds to a similar reduction in essential TB services.

It should be noted that these 26 high-burden countries account for approximately 80% of the Global TB burden and 90% of the TB burden within the Global Fund’s TB portfolio (and, consequently, among countries dependent on foreign aid for delivering TB services).

## Discussion

With TB services recovering to pre-COVID-19 levels and most excess cases from pandemic-related disruptions now identified and treated, a new, potentially greater threat to TB SDG targets has emerged. The freeze and cut in U.S. funding has caused uncertainty within the global TB community, the ambitious goals outlined in the End TB Strategy and the operational framework set out in the TB Global Plan 2023-2030, is under threat. The Global Plan highlights the critical role of new tools, including the development and rollout of a large-scale vaccine for TB, in achieving the End-TB targets. As a key funder for TB research, the U.S. withdrawal threatens the development of such critical new tools, further delaying progress and making End TB targets nearly unattainable. Hard-to-reach and high-risk and vulnerable populations, previously connected to national TB programs through civil society organizations (CSOs), now face complete isolation as U.S.-funded initiatives shut down.

Pre-existing funding gaps have now widened, worsening resource shortages needed to combat TB. Result suggests that the scale of increase is most pronounced in the worst-case scenario (S3), particularly in high-dependency countries.

Our analysis provides a conceptual illustration, with several areas requiring further, more fine-grained analysis to improve on. A key limitation is the estimation of the U.S. contribution to total TB expenditure and the assumption that its withdrawal leads to a proportional reduction in essential TB services. We estimated the proportion of TB expenditure funded by the U.S. using published data from the WHO’s datasets.^5^ However, the reliability of these 2023 proportions to approximate proportions in future years remains uncertain, as does the extent to which they directly support essential TB services, as assumed.

To mitigate this, in finding global impact, we incorporated a lower-bound estimate by varying the impact category assigned to each country. However, even this conservative bound under a permanent disruption scenario remains highly concerning.

The WHO database for TB expenditure, which we used to estimate USAID’s contribution, includes Global Fund (GF) sources (to which USAID contributes one- third) but captures only about 50% of the non-GF congressional appropriation for TB. We did not incorporate the remaining 50% in our analysis due to a lack of detailed information on its allocation across countries and its contribution to essential TB services. As a result, any potential overestimation that arises from our assumption that a proportional funding reduction leads to an equivalent reduction in essential TB services is partially offset by this omission.

Additionally, in optimistic scenarios where countries recover within three months or one year, they may shift to a low-dependency category - a process not accounted for in our future and medium impact estimates.

This leads to a broader point, which is not explored in this short- and medium-term impact analysis. The abrupt discontinuation of aid, no matter if or how it may be eventually restored, may force many countries to rethink its dependency not just US but on all foreign aid, and invest in new strategic planning for key health areas, not just TB. While the medium-term impact we studied here is alarming, the long-term picture may be more optimistic with countries becoming more self-reliant and more resilient to disruptions.

Our analysis shows that across 26 high-burden countries which account for 80% of the global TB burden and 90% among recipients of US funding for TB, termination of US funding will result in an estimated 10.6 million additional TB cases and 2.2 million additional TB deaths during the period 2025 to 2030. Many countries now stand at a crossroads: one path leads to continued reliance of foreign aid, while the other moves toward self-sufficiency and self-reliance.

## Contributors

SS, LD conceived the study. SM and CP developed the model and performed the analysis. SM, SN, SS, LD and CP all contributed to the interpretation of the results. SM and CP wrote a first draft of the manuscript; all authors contributed to developing the final draft. All authors read and approved the final manuscript.

## Supporting information

Technical Appendix

## Data Availability

We used publicly available data from the WHO website.

https://www.who.int/teams/global-tuberculosis-programme/data

## Acknowledgements

SM and CP are supported by the Avenir Health, USA. SN, SS and LD are supported by the Stop TB Partnership, Switzerland.

