## Supplementary material for "A Deadly Equation: The Global Toll of US TB Funding Cuts": Technical Appendix

**U.S. Funding for Global Tuberculosis program**

USAID was the leading bilateral donor in the global fight against tuberculosis, contributing approximately one third of international donor funds dedicated to TB control. This robust financial backing is complemented by annual congressional appropriations for TB control, which are expected to reach around 406 million USD in 2024. Such a significant investment underscores the United States' commitment to advancing TB diagnosis, treatment, and prevention efforts worldwide, reinforcing its pivotal role in the global initiative to curb this persistent public health challenge. Figure S1 depicts the annual TB funding of USAID since 2015 to 2024. Average annual increase of funding during this period is around 6%.

U.S. Funding for Global Tuberculosis (TB), FY 2015 - FY 2024 (USD millions)

|  |  |
| --- | --- |
| 2015 | 242 |
| 2016 | 240 |
| 2017 | 243 |
| 2018 | 272 |
| 2019 | 312 |
| 2020 | 317 |
| 2021 | 328 |
| 2022 | 380 |
| 2023 | 406 |
| 2024 | 406 |

**Figure S1**: Annual US congressional appropriations for TB

**Table S1: Country Classification Based on USAID Funding for the Tuberculosis Program**

| Low Dependency (0%–22%) | | Moderate Dependency (23%–37%) | | High Dependency (>37%) | |
| --- | --- | --- | --- | --- | --- |
| **Country (8)** | **Dependency on USAID** | **Country (10)** | **Dependency on USAID** | **Country (8)** | **Dependency on USAID** |
| India | 15.0% | Bangladesh | 36.0% | Mozambique | 45.0% |
| Peru | 1.3% | Pakistan | 25.3% | Myanmar | 37.9% |
| Thailand | 5.9% | Philippines | 26.3% | Nigeria | 40.3% |
| Angola | 8.7% | Viet Nam | 27.6% | Uganda | 41.3% |
| Nepal | 12.9% | Ghana | 29.7% | Ethiopia | 42.4% |
| Ukraine | 16.6% | Cameroon | 30.9% | Afghanistan | 44.1% |
| South Africa | 21.1% | Madagascar | 33.0% | Congo (Democratic Republic) | 47.3% |
| Indonesia | 22.5% | Zambia | 33.5% | Zimbabwe | 55.0% |
|  |  | Kenya | 34.3% |  |  |
|  |  | Tanzania (United Republic) | 35.8% |  |  |

**Model overview**

To estimate the potential impact of U.S. fund freezing on TB epidemic, we developed a deterministic compartmental model of tuberculosis (TB) transmission, illustrated in Figure S2. The model captures essential features of natural history of TB, like latent infection, asymptomatic, symptomatic stages as well as it captures country specific health care system. The disease compartments are also stratified with drug sensitive and drug-resistant strains. All model parameters listed in Table S3.

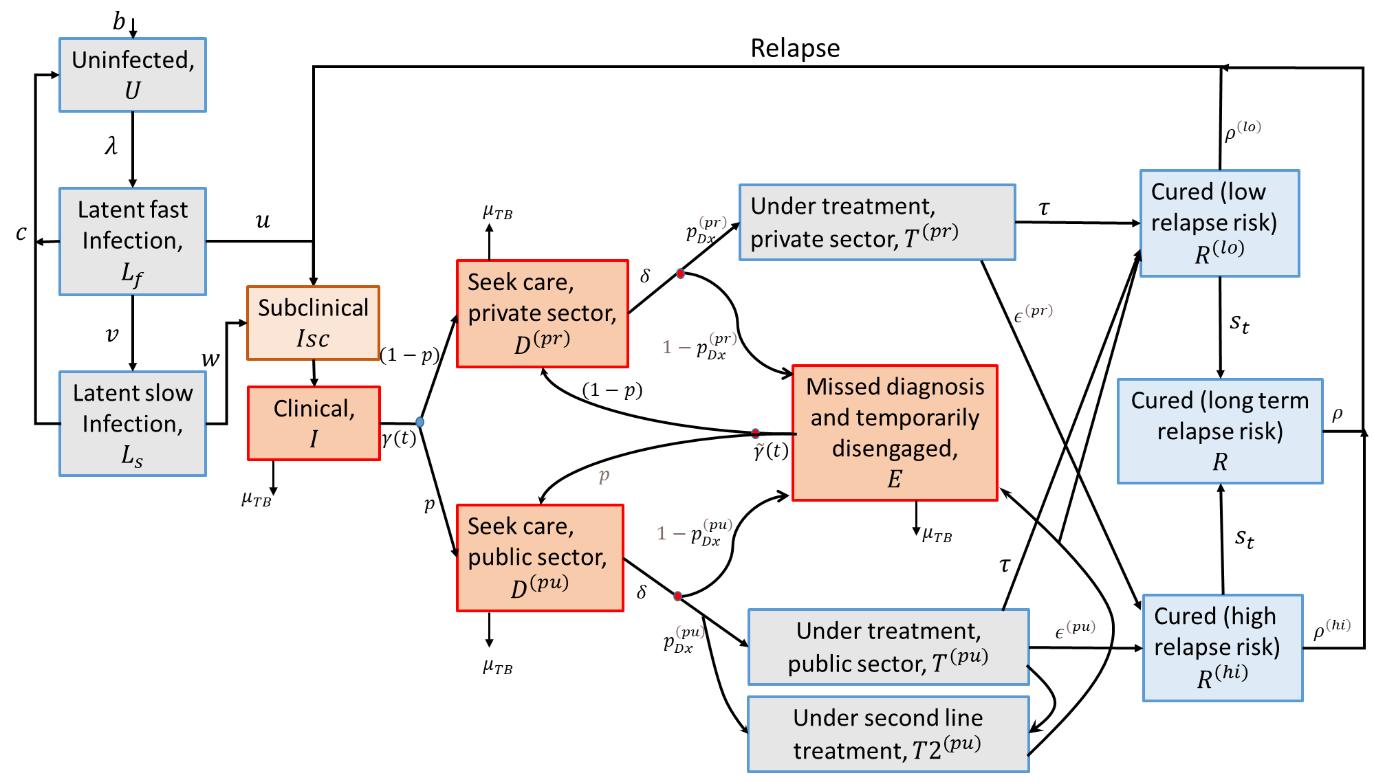

**Figure S2. Schematic diagram of the model structure.** Parameters are defined in Table S3. Infectious compartments contributing the force-of-infection are shown in red. For clarity, the diagram omits certain rates incorporated in the model, including self-cure; exogenous reinfection; and background mortality.

**Model equations**

The governing equations of the model are as follows. All state variables are written as proportions of the population (not as absolute numbers).

Uninfected (U):

$$\frac{dU}{dt}=b+c \sum_{s} \left( L_{s}^{\left( fast \right)}+L_{s}^{\left( slow \right)} \right)-U\sum_{s} \lambda_{s}- \mu U$$

for a birth-rate $b$; force-of-infection $\lambda_{s}$; s denotes the infecting strain (denoting drug-susceptible (s = 0) and drug-resistant (s = 1) TB); rate of clearance of LTBI $c$; and background mortality rate $\mu$.

Latent, ‘fast’ infection ($L_{s}^{(fast)}$):

$$\frac{dL_{s}^{\left( fast \right)}}{dt}=\lambda_{s}\left[ U+\left( 1-h \right) \left( R_{s}^{(hi)}+R_{s}^{(lo)}+R_{s}^{(st)} \right) \right]-\left( \mu+u+v+c \right)L_{s}^{\left( fast \right)}$$

for a progression rate $u$; a ‘stabilisation’ rate (to latent ‘slow’ status) $v$; and protection from reinfection $h$, amongst those previously infected.

Latent, ‘slow’ infection ($L_{s}^{\left( slow \right)}$):

$$\frac{dL_{s}^{\left( slow \right)}}{dt}=v L_{s}^{\left( fast \right)}-\left( \mu+c+w \right)L_{s}^{\left( slow \right)}$$

for a reactivation rate $w$.

Subclinical TB ($I_{s}^{\left( sc \right)}$):

$$\frac{dI_{s}^{\left( sc \right)}}{dt}=u L_{s}^{(fast)}+wL_{s}^{\left( slow \right)}+\rho^{(hi)}R_{s}^{(hi)}+\rho^{(lo)}R_{s}^{(lo)}+\rho^{(st)}R_{s}^{(st)}-\left( \mu+\sigma+r \right)I_{s}^{\left( sc \right)}$$

for relapse rates $\rho^{\left( hi \right)}, \rho^{\left( lo \right)}, \rho^{\left( st \right)}$; rate of developing clinical symptom $r$ and self-cure rate $\sigma$.

Clinical TB ($I_{s}$):

$$\frac{dI_{s}}{dt}=r I_{s}^{\left( sc \right)}-(\gamma+\mu^{(TB)}+\sigma)I_{s}$$

for care-seeking rate $\gamma$ and TB mortality rate $\mu^{(TB)}$.

Presented for diagnosis with provider type $j$, ($D_{s}^{(j)}$):

$$\frac{dD_{s}^{(pu)}}{dt}= p (\gamma I_{s}+ \tilde{\gamma} E_{s})-\left( \mu^{(TB)}+\sigma+\delta\right)D_{s}^{(pu)}$$

$$\frac{dD_{s}^{(pr)}}{dt}=(1-p) (\gamma I_{s}+ \tilde{\gamma} E_{s})-\left( \mu^{(TB)}+\sigma+\delta\right)D_{s}^{(pr)}$$

Here, $p$ and $(1-p)$ are, respectively, the proportion-of-presentation to healthcare providers in the public and private sectors. Rate-of-offering diagnosis $\delta$. Here,  $\tilde{\gamma}$is analogous to $\gamma$but attached to individuals who remain undiagnosed despite having previously sought care (i.e. compartment $E,$below).

On TB treatment with provider type $po,$ with DS TB ($T_{0}^{(po)}$) initiating first line treatment

$$\frac{dT_{0}^{(pu)}}{dt}=\delta p_{Dx}^{\left( pu \right)}\left( D_{0}^{(pu)}+(1-{DST}_{(+ve)})D_{1}^{(pu)} \right)-\left( \mu+\tau+\epsilon^{\left( pu \right)}+MDR\_acqu \right)T_{0}^{(pu)}$$

$$\frac{dT_{0}^{(pr)}}{dt}=\delta p_{Dx}^{\left( pr \right)}D_{0}^{(pr)}-\left( \mu+\tau+\epsilon^{\left( pr \right)}+MDR\_acqu \right)T_{0}^{(pr)}$$

for a treatment completion rate $\tau$; and a treatment interruption rate $\epsilon^{\left( po \right)}$. Acquired MDR-TB during first-line treatment in the respective sectors,

$$\frac{dT_{1}^{(pu)}}{dt}=\left( MDR\_acqu \right)T_{0}^{(pu)}$$

$$\frac{dT_{1}^{(pr)}}{dt}=\left( MDR\_acqu \right)T_{0}^{(pr)}$$

Drug resistance TB initiating second line treatment ($S_{1}^{(pu)}$)

$$\frac{dS_{1}^{(pu)}}{dt}=\delta p_{Dx}^{\left( pu \right)} {DST}_{(+ve)} D_{1}^{(pu)}+SLtrans*T_{0}^{(pu)}-\left( \mu+T+{\epsilon2}^{\left( pu \right)} \right)T_{0}^{(pu)}$$

For treatment completion rate $T$; and a treatment interruption rate ${\epsilon2}^{\left( po \right)}$. SLtrans represents transfer to SL treatment during FL treatment.

Missed diagnosis and temporarily disengaged from care-seeking ($E_{s}$):

$$\frac{dE_{s}}{dt}=\delta\left( 1-p_{Dx}^{\left( pu \right)} \right)D_{s}^{(pu)}+\delta\left( 1-p_{Dx}^{\left( pr \right)} \right)D_{s}^{(pr)}-\left( \mu+\sigma+\tilde{\gamma} \right)E_{s}$$

Recovered with low relapse risk, following treatment completion ($R^{(lo)}$):

$$\frac{dR_{s}^{(lo)}}{dt}=\tau\left( T_{s}^{(pu)}+T_{s}^{(pr)} \right)-[(1-h)\lambda_{s}+\rho^{\left( lo \right)}+\mu+s_{t}]R_{s}^{(lo)}$$

for a rate of ‘stabilisation’ of relapse risk $s_{t}$.

Recovered with high relapse risk, following treatment completion ($R_{s}^{(hi)}$):

$$\frac{dR_{s}^{(hi)}}{dt}=\left( \epsilon^{\left( pu \right)} T_{s}^{(pu)}+\epsilon^{\left( pr \right)}T_{s}^{(pr)} \right)-[(1-h)\sum_{s} \lambda_{s}+\rho^{\left( hi \right)}+\mu+s_{t}]R_{s}^{(hi)}$$

Long-term, ‘stabilised’ relapse risk ($R_{s}^{(st)}$):

$$\frac{dR_{s}^{(st)}}{dt}=s_{t}\left( R_{s}^{(lo)}+R_{s}^{(hi)} \right)-\left[ \left( 1-h \right)\lambda_{s}+\rho^{\left( st \right)}+\mu\right]R_{s}^{\left( st \right)}+f\left( R_{s}^{(st)} \right)$$

Force-of-infection $(\lambda_{s})$:

$$\lambda_{0}=\sum_{h} \beta_{ds} ({k I}_{0}^{\left( sc \right)}+{I_{0}+E}_{0}+D_{0}^{(pu)}+D_{0}^{(pr)})$$

$$\lambda_{1}=\sum_{h} \beta_{dr} ({k I}_{1}^{\left( sc \right)}+{I_{1}+E}_{1}+D_{1}^{(pu)}+D_{1}^{(pr)})$$

Where $\beta_{ds}$ is the rate-of-transmission associated with drug-susceptible and $\beta_{dr}$ is associated with drug-resistance TB disease. $k$ is the infectiousness of subclinical TB relative to clinical TB.

**Model calibration**

We adjusted model parameters to match WHO estimates for incidence (all forms of TB) in 2023; TB induced mortality rate in 2023; TB notifications in 2023; incidence of RR/MDR-TB in 2019 and in 2023; second line treatment initiation in 2023 and prevalence of subclinical TB in 2023. Calibration targets for fitting the model with the data are summarized in Table S2.

**Table S2.** Calibration targets for the model countries^1^

| Indicators | Country 1 | Country 2 | Country 3 |
| --- | --- | --- | --- |
| Incidence per 100,000 population in 2023 | 195 [168 - 228] | 221 [161 - 291] | 361 [220 - 537] |
| Notification per 100,000 in 2023 | 175 [+/- 15%] | 177 [+/-15%] | 347 [+/-15%] |
| Mortality per 100,000 in 2023 | 22 [16 – 30] | 26 [15 – 38] | 13 [4.7 – 25] |
| MDR incidence per 100,000in 2019 | 8.2 [6.7 – 9.7] | 2.97 [1.5 – 4.6] | 13 [7.4 – 19] |
| MDR incidence per 100,000 in 2023 | 7.4 [5.7 – 9.1] | 2.9 [1.3 – 5.0] | 12 [6.9 – 18] |
| Second line treatment initiation per 100,000 in 2023 | 3.45 [+/- 15%] | 1.2 [+/- 15%] | 4.75 [+/- 15%] |
| Proportion of prevalent cases that are symptomatic. | 0.5 [0.36 – 0.8] | 0.5 [0.36 – 0.8] | 0.5 [0.36 – 0.8] |

Calibration was conducted using Bayesian Markov Chain Monte Carlo (MCMC).^2^ Specifically, we created likelihood functions for each calibration target, incorporating both central values and uncertainty intervals. A posterior density was then constructed by combining these likelihood functions with uniform priors for the model parameters. Sampling from this posterior density was performed using an adaptive algorithm8. After eliminating the burn-in phase and applying ‘thinning’, we obtained 250 samples for simulation. For all model projections, uncertainty was quantified by defining the interval between the 2.5th and 97.5th percentiles as the ‘95% Bayesian credible interval’. The central estimate was determined using the 50th percentile.

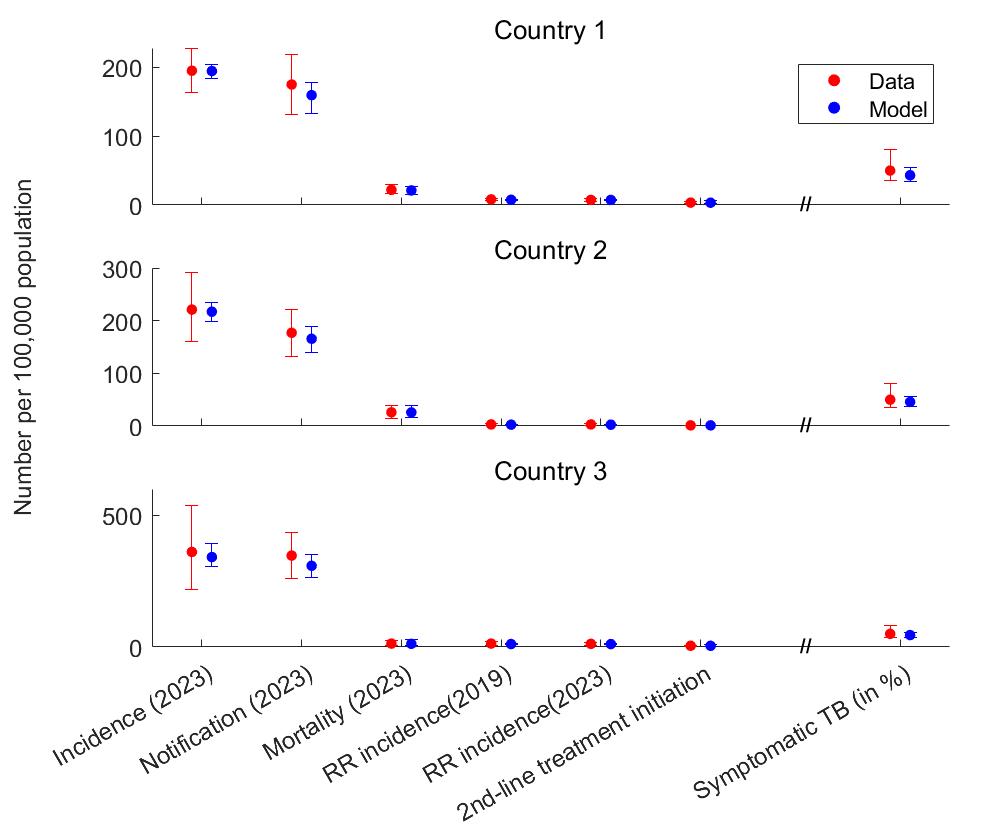

**Figure S3**. Model calibration with the observed data for the three countries. Dots show central estimates, while error bars show 95% uncertainty intervals.

**Modelling disruption due to fund freezing**

The primary service disruption expected from funding cuts is a reduction in case-finding activities and early TB diagnosis. We assumed that the rate-of-presentation to care (denoted as parameter $\gamma$) in the public sector would decline proportionally with reduced funding. Similarly, the probability of achieving successful TB diagnosis and initiating treatment per care-seeking event ($p_{Dx}^{\left( pu \right)}$) is anticipated to decrease proportionately. Additionally, funding disruptions could affect other stages of the program, depending on country-specific allocations, such as drug susceptibility testing, treatment for drug-resistant TB, and TB infection prevention interventions. For simplicity, our analysis focuses solely on the major case-finding activity, omitting these individual interventions.

These parameters are assumed to decrease in three-month time. This reduction is assumed to be proportional to the funding contribution of USAID on TB in that country. Three recovery scenarios have been developed to examine the impact of fund freezing.

a. Minimum Impact Scenario (S1): the country recovers service coverage to baseline levels within next three months after disruption.

b. Moderate Impact Scenario (S2): it takes one year for the country to recover service coverage to baseline levels.

c. Worst Impact Scenario (S3): The service coverage remains at a lower level permanently.

In all these scenarios scale-down or scale-up occurred linearly as per mentioned period.

**Table of parameters**

**Table S3:** **List of model parameters**: Parameters and their symbol as depicted in figure 2S and in the model equations. The labels ‘C1,’ ‘C2,’ and ‘C3’ indicate parameter values specific to Country 1, Country 2, and Country 3, respectively.

| Parameter | | Symbol | Values | Note/references |
| --- | --- | --- | --- | --- |
| Natural history | | | |  |
| Infection rate (number of annual infections per case) | | $\beta$ | 25 [13 - 35]; C1  30 [18 - 39]; C2  31 [16 - 39]; C3 | Estimated |
| Infection rate for drug resistance TB relative to drug susceptible TB | | $\beta_{mdr}$ | 0.34 [0.23 – 0.44]; C1  0.26[0.11 – 0.51]; C2  0.28[0.18 – 0.37]; C3 | Estimated |
| Per-capita annual rate of progression from ‘fast’ latent infection | | $u$ | 0.078 [0.065 – 0.100]; C1  0.085 [0.063 – 0.103]; C2  0.083 [0.063 – 0.102]; C3 | Estimated using Menzies (2018)^3^ |
| Per-capita annual rate of stabilisation from ‘fast’ to ‘slow’ latent status | | $v$ | 0.872 | Menzies (2018) ^3^ |
| Per-capita annual rate of reactivation from ‘slow’ latent infection | | $w$ | 0.0006 [0.0004 – 0.0007]; C1  0.0006 [0.0004–0.0007]; C2  0.0006 [0.0004–0.0007]; C3 | Menzies (2018)^3^ |
| Per-capita annual rate of self-clearance of latent TB | | $c$ | 0.031 (0.022 – 0.035) | Emery (2021)^4^ for central value, and taking uniform priors of +/- 25% |
| Per-capita annual rate of developing symptoms, amongst subclinical TB | | $r$ | 56 [12 – 98]; C1  54 [12 – 97]; C2  51 [9 – 94]; C3 | Estimated |
| Per-capita annual rate of TB mortality while untreated | | $\mu_{TB}$(general population) | 0.22 [0.12 – 0.35]; C1  0.31 [0.15 – 0.49]; C2  0.07 [0.02 – 0.18]; C3 | Estimated |
| Protection from reinfection amongst those with prior infection | | $h$ | 0.52 [0.39 – 0.61]; C1  0.51 [0.38 – 0.61]; C2  0.47 [0.38 – 0.59]; C3 |  |
| Per-capita annual rate of relapse in first two years after treatment completion | | $\rho^{\left( lo \right)}$ | 0.08 [0.065 – 0.105]; C1  0.08 [0.064 – 0.104]; C2  0.08 [0.064 – 0.105]; C3 | Thomas A et al (2005)^5^, Romanowski (2019)^6^, Menzies (2009)^7^ and Weis (1994)^8^, with uniform prior using intervals of ± 5% |
| Per-capita annual rate of relapse in first two years after self-cure or incomplete treatment | | $\rho^{\left( hi \right)}$ | 0.139 [0.110 – 0.170]; C1  0.140 [0.107 – 0.174]; C2  0.117 [0.106 – 0.147]; C3 |  |
| Per-capita annual rate of relapse>two years after last TB episode | | $\rho$ | 0.0014 [0.0011 – 0.0018]; C1  0.0016 [0.0012– 0.0019]; C2  0.0015 [0.0011– 0.0019]; C3 | Most relapse occurs in first two years after recovery: Guerra-Assuncao (2015)^9^ |
| Per-capita annual rate of ‘stabilising’ from high to low relapse risk | | $s$ | 0.5 y^-1^ | Assumed |
| Rate-of-presentation to care, first care-seeking visit | | $\gamma$ | 2.13 y^-1^ [1.2 – 3.31]; C1  2.29 y^-1^ [1.4 – 3.44]; C2  2.17 y^-1^ [1.4 – 3.19]; C3 | Estimated |
| Rate-of-presentation to care, second and subsequent care-seeking visits | | $\tilde{\gamma}$ | 9.5 y^-1^ [1.3 – 28.3]; C1  58 y^-1^ [7.3 – 135]; C2  43 y^-1^ [3.3 – 126]; C3 | Estimated |
| Probability that a TB patient visits public provider, per care-seeking attempt | | $p$ | 0.83 [0.58 – 0.99]; C1  0.84 [0.47 – 0.99]; C2  0.92 [0.70 – 0.99]; C3 | Estimated |
| Per-capita rate of offering diagnosis | | $\delta$ | 52 y-1 | Assumption: corresponds to an average of 1 week to arrive at a diagnosis |
| Probability of successful TB diagnosis and treatment initiation per care-seeking visit | Public sector | $p_{Dx}^{\left( pu \right)}$ | 0.83 [0.75 – 0.89]; C1  0.81 [0.75 – 0.88]; C2  0.83 [0.75 – 0.89]; C3 | Estimated |
|  | Private sector | $p_{Dx}^{\left( pr \right)}$ | 0.25 [0.10 – 0.65]; C1  0.30 [0.10 – 0.65]; C2  0.34 [0.10 – 0.67]; C3 | Estimated |
| Per-capita annual rate of TB self-cure | | $\sigma$ | 0.17 [0.12 – 0.20]; C1  0.16 [0.12 – 0.20]; C2  0.17 [0.13 – 0.21]; C3 | Tiemersma et al., (2011)^10^ for central value, with uniform prior using intervals of ± 15% |
| Of diagnosed TB with rifampicin resistance, proportion that is recognized as such in 2019 (through DST) | | ${DST}_{(+ve)}$ | 0.23 [0.12 – 0.47]; C1  0.20 [0.11 – 0.34]; C2  0.18 [0.11 – 0.28]; C3 | Calibrated for simulated, second-line treatment initiations to match reported RR/MDR notifications |
| Per capita rate of acquired RR/MDR during treatment | | MDR_acqu | 0.04 [0.02 – 0.06]; C1  0.03 [0.003 – 0.06]; C2  0.04 [0.01 – 0.06]; C3 | Estimated |
| Rate of transfer to SL treatment during FL treatment. | | $SLtrans$ | 0.88 | Assumption |
| Per-capita annual rate of treatment completion | | $\tau$ | 2 y-1 (for Firstline treatment)  0.5 y-1 (for Second line treatment) | Corresponds to a 6-month regimen  Corresponds to a 2-year regimen |
| Per-capita annual rate of treatment interruption | Public sector | $\epsilon^{\left( pu \right)}$ | 0.29 [0.11 – 0.64]; C1  0.38 [0.11 – 0.62]; C2  0.50 [0.18 – 0.66]; C3 | Calculated using $\epsilon^{\left( pu \right)}=\frac{1-P}{P}\tau$, for treatment completion rate $P$, and assuming U[0.75, 0.95] for $P$ |
|  | Private sector | $\epsilon^{\left( pr \right)}$ | 1.5 [0.56 – 2.81]; C1  1.6 [0.57 – 2.83]; C2  1.1 [0.53 – 2.77]; C3 |  |
| Demographics | | | |  |
| Per-capita annual rate of background mortality | | $\mu$ (general population) | 1/68 y-1; C1  1/74 y-1; C2  1/60 y-1; C3 | Corresponds to average lifespan (World Bank 2022)^11^ |

**Results**

Figure S4 shows the impact of US fund cut in Country 1, Country 2 and Country 3.

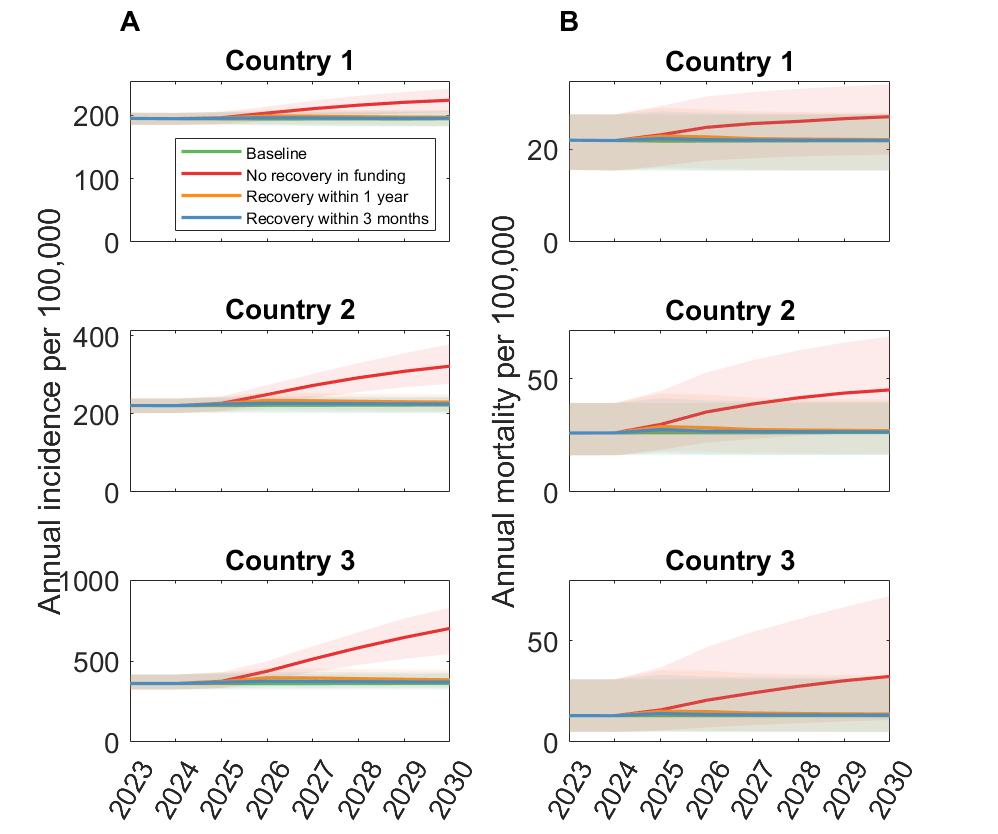

**FigureS4: Impact of a US Aid freeze on tuberculosis outcomes across three dependency categories: Country 1(low dependency), Country 2 (moderate dependency), and Country 3 (high dependency).** (A) Annual TB incidence per 100,000 population under three funding scenarios: a 90-day freeze with a subsequent 3-month recovery, a one-year recovery, and permanent disruption (no recovery). (B) Annual TB mortality rate corresponding to the same three recovery scenarios.

**Table S4:** Impact of temporary and permanent disruption resulting from US funding freeze for TB

|  | **uninterrupted scenario (S0)** | **Minimum impact scenario (S2)** | **Moderate impact scenario (S2)** | **Worst impact scenario (S3)** |
| --- | --- | --- | --- | --- |
|  | Baseline | 90-day disruption, 90- day recovery | 90-day disruption, 1 year recovery | permanent disruption |
| **New TB cases** |  |  |  |  |
| Total for 2025-2030 | 46,679,800 | 47,280,400 | 48,254,900 | 56,653,200 |
| Additional new TB cases relative to baseline |  | 600,600 [420,900-796,900] | 1,575,100 [1,146,300-2,058,600] | 9,973,400 [7,267,400-13,144,000] |
| Annual increase |  | 100,100 [70,200-132,800] | 262,500 [191,100-343,100] | 1,662,200 [1,211,200-2,190,700] |
| **TB Deaths** |  |  |  |  |
| Total for 2025-2030 | 6,678,800 | 6,787,100 | 6,968,900 | 9,119,500 |
| Additional TB deaths relative to baseline |  | 108,300 [74,500-144,000] | 290,100 [211,100-379,100] | 2,440,700 [1,778,500-3,216,600] |
| Annual increase |  | 18,100 [12,400-24,000] | 48,400 [35,200-63,200] | 406,800 [296,400-536,100] |
| **US funding for TB 2025-2030** |  | **Note** |  |  |
| Congressional appropriation | $ 2,394,671,440 | approximately 80% of the appropriation to these HB countries | |  |
| GF contribution | $ 1,264,000,000 | ~632 million USD to these HB countries, 1/3 from US funding | |  |
| Total | **$ 3,658,671,440** |  |  |  |

**Data sharing**

The model code and dataset are publicly available at (we’ll provide GitHub link)
